# Expression profiles of circRNAs, lncRNAs, and mRNAs in extreme phenotypes of diabetic retinopathy

**DOI:** 10.1101/2020.02.15.20023481

**Authors:** Rouxi Zhou, Sen Liu, Wei Wang, Weijing Cheng, Miao He, Kun Xiong, Xia Gong, Yuting Li, Wenyong Huang

## Abstract

Recent evidences highlighted regulatory role of circular RNAs (circRNAs) and long non-coding RNAs (lncRNAs) in the development of diabetic retinopathy (DR). However, the literatures and number of the RNAs identified were limited. Here, we compared the expression profiles of circRNAs, lncRNAs, and mRNAs in the blood of the susceptible individuals who developed severe DR within 5 years after being diagnosed with type 2 diabetic mellitus (T2DM), and the inherently resistant individuals who are spared from DR despite over 20-year history of T2DM. Using RNA microarray, hundreds of significantly differently expressed circRNAs, lncRNAs, and dozens of mRNAs were identified. Quantitative polymerase chain reaction verified the above findings. Gene ontology analysis indicated that the differentially expressed circRNAs were involved in platelet-derived growth factor binding, and mRNA and the cis-target genes of lncRNA participate in negative regulation of the Wnt signaling pathway. Kyoto Encyclopedia of Genes and Genomes pathway analysis suggested that the differentially expressed circRNAs were related to vitamin B6 metabolism and type 2 diabetes. The cis-target genes of lncRNAs are enriched in valine, leucine, and isoleucine biosynthesis and in the hypoxia-inducible factor-1 signaling pathway. The trans-target genes of lncRNAs are enriched in pathways such as vitamin B6 metabolism. Differentially expressed mRNAs are associated with type 2 diabetes and the vascular endothelial growth factor signaling pathway. Our findings demonstrate that circRNAs and lncRNAs may be involved in the regulation of DR and lay a foundation for further researches into the underlying mechanisms.

## Introduction

Diabetic retinopathy (DR) is one of major microvascular complications of diabetic mellitus (DM), as well as the leading cause of blindness for working-age population[1]. It is estimated that approximately one third of DM patients are suffering from DR and 80% patients with a history of DM more than 20 years will develop DR[2, 3]. DR has become an important global public health problem as the number of DM patients explodes and continues to explode in recent years[4, 5]. The cause of development and progression of DR is still not well-understood. Duration of diabetes, hemoglobin A1c, cholesterol and blood pressure only accounted for approximately 10% of the risk of developing retinopathy[6, 7]. Genetics contributes to about 27% of the susceptibility of development of DR[8]. The above factors are far from adequate, implying the possible role of epigenetics in the pathogenesis of DR.

Non-coding RNAs (ncRNAs) play an essential part in epigenetic regulation, among which circular RNAs (circRNAs) and long non-coding RNAs (lncRNAs) have attracted considerable attention in recent years [9, 10]. Circular RNAs (circRNAs) are a group of non-coding RNAs that have the covalently closed loop structure. They are widely found in eukaryotes, and they regulate gene expression at the transcriptional and post-transcriptional levels [11]. Over the past few years, the role of circRNAs in DR has been brought into focus. We and other researchers have found changes in the expression profiles of circRNAs in the blood, vitreous, and retina of patients with DR [12-14]. LncRNAs are non-coding RNAs longer than 200bp, and involved in numerous biological processes and participate in almost all aspects of gene expression, from transcription to mRNA splicing, RNA decay, and translation [15]. Recent studies have revealed that DR is characterized by abnormal expression of lncRNAs, and lncRNAs are involved in the pathogenesis of DR [16, 17]. Although recent findings have unraveled significance of circRNAs and lncRNAs, the literature and number of RNAs identified are limited, and the effects of the vast majority of circRNAs and lncRNAs remain unknown.

The population in the Pearl River Delta in China is approximately 100 million. However, the epigenetic and genetic epidemiological studies of DR in this area are still lacking to date. We have built a cohort of type 2 DM (T2DM) patients in this area, Guangzhou Diabetic Eye Study (GDES) cohort, to investigate the genetic, epigenetic and environmental risk factors for the development of DR. In the present study, we selected individuals with extreme phenotypes from the cohort, and compare the expression profiles of circRNAs, lncRNAs and mRNAs to explore role of the ncRNAs in the pathogenesis of DR.

## Materials and Methods

### Patients and samples

The patients were enrolled from the GDES, the largest cohort study including 2305 T2DM patients in southern China. The study protocol was approved by the ethical review committee of Zhongshan Ophthalmic Center and conducted in accordance with the tenets of the Helsinki Declaration. Written informed consent was obtained from each subject before enrollment.

The methodology of GDES was described in detail in our other report. In brief, GDES recruited patients diagnosed with type 2 DM between the age of 35 and 85 and having no prior ocular treatment. Patients underwent comprehensive ophthalmic examinations, including visual acuity assessment, intraocular pressure (IOP) measurement (Topcon CT-80A non-contact tonometry; Topcon, Japan), standardized 7-field 45° colour retinal photographs (Canon CX-1, Tokyo, Japan), optical coherence tomography (Heidelberg Spectralis; Heidelberg Engineering, Heidelberg, Germany), optical coherence tomography angiography (Triton DRI-OCT, Topcon. Inc., Tokyo, Japan), blood and urine sample collection.

Patients exhibited the following extreme phenotypes were selected from the cohort and included for this analysis: (1) group 1 (control): patients who had a history of more than 20 years of DM but no retinopathy; (2) group 2 (DR): patients who developed DR and diabetic macular edema (DME) within 5 years after being diagnosed with DM. The diagnosis and classification of DR was based on color retinal photograph by the Early Treatment Diabetic Retinopathy Study (ETDRS) criteria[18].

### RNA extraction

All blood samples were sent to SHBIO (Shanghai, China) for RNA analysis. PX Blood RNA Kit (Cat # R1057-02, Omega Bio-tek, Inc., GA, USA) was used for the purpose, and total RNAs were extracted from the blood samples according to the standard operating procedures specified by the kit manufacturer. The initial sample in the microarray was composed of total RNAs, and these total RNAs were tested using a NanoDrop ND-2000 spectrophotometer (Thermo Fisher Scientific, Inc., Waltham, MA, USA) and an Agilent Bioanalyzer 2100 (Agilent Technologies, Santa Clara, CA, USA). The qualified RNA was used in the subsequent experiments.

### RNA microarray

RNA-expression-profiling microarray operations were performed by following quasi-operational procedures. The total RNAs were purified using a RNeasy mini kit (Cat. # 74106, Qiagen, GmBH, Germany). They were amplified and labeled using the One-color Low Input Quick Amp Labeling Kit (Cat. # 5190-2305, Agilent Technologies, Santa Clara, CA, USA) by following the manufacturer’s instructions. Each slide was hybridized with 600 ng Cy3-labeled cRNA by using a Gene Expression Hybridization Kit (Cat. # 5188-5242, Agilent Technologies, Santa Clara, CA, USA) in a Hybridization Oven (Cat. # G2545A, Agilent Technologies, Santa Clara, CA, USA) by following the manufacturer’s instructions. After 17 h of hybridization, the slides were washed in staining dishes (Cat. # 121, Thermo Shandon, Waltham, MA, USA) with a Gene Expression Wash Buffer Kit (Cat. # 5188-5327, Agilent Technologies, Santa Clara, CA, USA). The washed slides were scanned using an Agilent Microarray Scanner (Cat. # G2565CA, Agilent Technologies, Santa Clara, CA, USA) with the default settings. The scanning parameters were as follows: Dye channel: green; Scan resolution = 3 μm; PMT: 100%; 20 bits. Data were extracted using Feature Extraction software 10.7 (Agilent Technologies, Santa Clara, CA, USA). The raw data were normalized using the quantile algorithm included in the limma package in R.

### qPCR verification

The cDNA was placed in the QuantStudio 5 Real-Time PCR System (Thermo Fisher Scientific, Inc., Waltham, MA, USA), which was operated using the QuantStudio™ Design and Analysis Software (Thermo Fisher Scientific, Inc., Waltham, MA, USA). The procedure was as follows: 50 °C, 2 min; 95 °C, 10 min (95 °C, 15 s; 60 °C, 1 min) for 40 cycles. The primers corresponding to each circRNA are listed in Table 1.

**Table 1.**
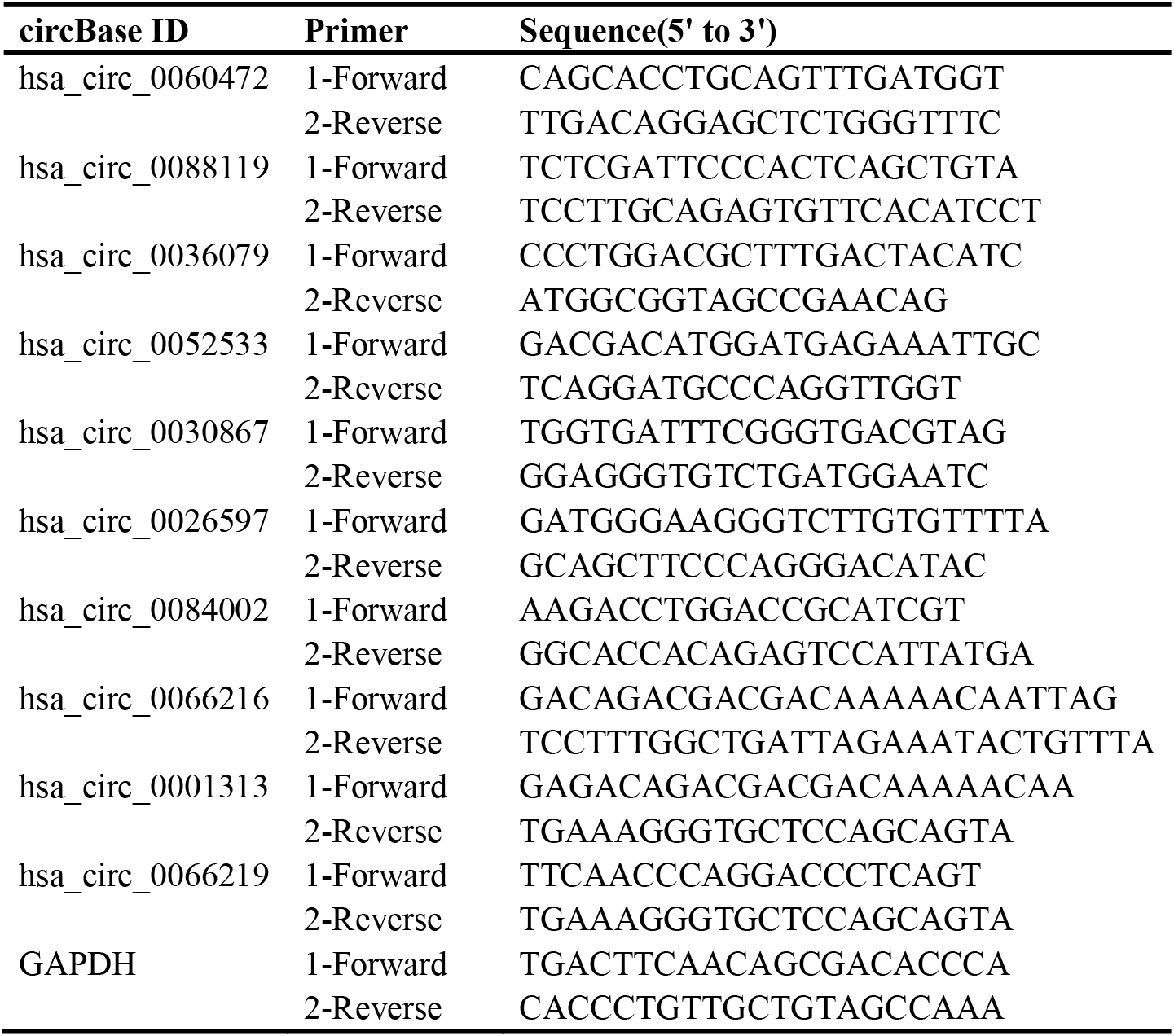
Primers used for qPCR validation of randomly selected circRNAs.

### Screening of differentially expressed RNAs

Normalization of raw data: The raw data obtained in the scan were normalized using the quantile algorithm included in the limma package in R. The normalized signal value was the signal value calculated using log2. Screening of differential RNAs: After data normalization, the differential RNAs were screened using fold change (multiple expression differences) and Student’s t-test. The criteria for significant difference were as follows: (1) fold change (linear) ≤ 0.5 or fold change (linear) ≥ 2.0; (2) t-test P < 0.05. A heat map was used to illustrate the expression levels of various RNAs in each sample. A scatter plot was created to evaluate the overall distribution of the two datasets. A volcano plot was generated to illustrate the number and distribution of the differentially expressed RNAs.

### GO and KEGG enrichment analysis of differentially expressed circRNAs, lncRNAs, and mRNAs

Gene Ontology (GO) is an international standard annotation system for the functions of genes and gene products. GO classifies functions into three domains: molecular function, biological processes, and cellular components [19-21]. Fisher’s exact test was used for enrichment analysis. The clusterProfiler (from R) and Bioconductor data analysis packages were used for the purpose. P < 0.05 was set as the significance level for a GO term. Kyoto Encyclopedia of Genes and Genomes (KEGG) pathway analysis was performed to identify the biological functions of the differentially expressed RNAs by mapping the RNAs onto the KEGG pathway map [22, 23]. P < 0.05 was considered to represent significant enrichment.

### Prediction of lncRNA target genes

A gene within 10 kb from a lncRNA on the same chromosome was selected as the target gene for testing the cis-regulatory effect of lncRNAs. BLASTN searches were conducted to find sequences that were complementary or similar to lncRNAs; then, RNAplex software was used to calculate the complementary energy between the two sequences, where sequences with e ≤ −30 were considered trans-target genes.

### CircRNA-microRNA interaction analysis

miRDB (http://www.mirdb.org) and TargetScan (http://www.targetscan.org/vert_72/) were used to predict the binding of microRNAs to circRNAs.

## Results

### Demographic and clinical characteristics of study participants

Four patients, including 3 males and 1 female were included in each group respectively. In group 1, there was no evidence of retinal and macular lesions in both eyes of all the patients. In group 2, 3 patients had moderate non-proliferative DR (NPDR) and 1 had severe NPDR in the right eye, among whom 3 had DME; in the left eye, 3 patients had moderate NPDR, 1 had severe NPDR, and DME occurred in all four patients.

As summarized in table 2, patients in group 1 have a mean age of 63.5±2.6 years, with an average 26.0±2.9 years of DM, while in group 2, the mean age is 58.8±8.3 years (P=0.316) and the average duration of DM is 3.0±1.2 years (P<0.001). The BMI, SBP, DBP, HbA1c level, triglycerides, cholesterol, high density lipoprotein cholesterol (HDL-c), low density lipoprotein cholesterol (LDL-c), creatinine, microalbuminuria (MAU), CRP (C-reactive protein), IOP and axial length were not significantly different between the two groups (all P>0.05). Central retinal thickness was significantly greater in patients in group 2 (272.3±17.7 vs. 303.4±0.8μm in the right eye, P=0.045; 193.3±21.5 vs. 321.5±33.2μm in the right eye, P=0.012) in comparisons with those in group 1.

**Table 2.** Demographic and clinical characteristics of study participants.

|  | **Group 1** | **Group 2** | **P-value** |
| --- | --- | --- | --- |
| Age, years | 63.5±2.6 (61–67) | 58.8±8.3 (49–69) | 0.316 |
| BMI, kg/m<sup>2</sup> | 25.0±2.8 (22.1–27.7) | 24.9±3.9 (21.5–30.5) | 0.985 |
| SBP, mmHg | 128.5±12.9 (116–145) | 135.3±27.5 (108–173) | 0.672 |
| DBP, mmHg | 70.5±14.5 (51–82) | 70.3±18.4 (45–88) | 0.984 |
| DM duration, year | 26.0±2.9 (23–29) | 3.0±1.2 (2–4) | **<0.001** |
| HbA1c level, % | 6.4±0.5 (6.1–7.1) | 7.4±0.8 (6.7–8.1) | 0.069 |
| Triglycerides, mmol/L | 1.6±0.6 (1.19–2.4) | 2.4±1.9 (1.1–5.18) | 0.448 |
| Cholesterol, mmol/L | 4.3±0.3 (4.07–4.69) | 4.5±1.4 (2.53–5.74) | 0.829 |
| LDL-c, mmol/L | 2.6±0.3 (2.36–2.97) | 3.0±1.3 (1.32–4.49) | 0.603 |
| HDL-c, mmol/L | 1.4±0.1 (1.19–1.45) | 1.0±0.4 (0.65–1.48) | 0.092 |
| Creatinine, µmol/L | 75.3±24.4 (50–104) | 81.0±18.6 (57–101) | 0.720 |
| MAU, mg/mL | 0.7±0.6 (0–1.24) | 6.7±9.1 (0–19.65) | 0.312 |
| CRP, mg/L | 0.4±0.2 (0.23–0.58) | 2.7±2.1 (1.35–5.75) | 0.123 |
| IOP, mmHg |  |  |  |
| Right eye | 18.2±3.2 (14.7–21.7) | 15.1±1.2 (13.3–16) | 0.123 |
| Left eye | 18.0±4.2 (14.3–24) | 14.7±1.2 (13.3–16) | 0.180 |
| Axial length, mm |  |  |  |
| Right eye | 23.1±0.7 (22.06–23.64) | 23.2±1.4 (21.11–24.23) | 0.945 |
| Left eye | 23.1±0.9 (21.79–23.62) | 23.2±1.4 (21.07–24.39) | 0.921 |
| Central RT, µm |  |  |  |
| Right eye | 272.3±17.7 (259–285) | 303.4±0.8 (303–304) | **0.045** |
| Left eye | 193.3±21.5 (179–218) | 321.5±33.2 (298–345) | **0.012** |

Data present as Mean ± SD (min to max).
DM=diabetes mellitus; RT= retinal thickness; MCT=macular choroidal thickness;
MAU=Microalbuminuria; IOP= Intraocular Pressure; HDL-c= High Density
Lipoprotein Cholesterol; LDL-c= Low Density Lipoprotein Cholesterol; CRP=
C-reactive protein.
Bold indicates statistical significance.

### Differential expression of circRNAs, lncRNAs, and mRNAs in DR and control

As shown in figure 1, a total of 494 circRNAs were differentially expressed between group 1 and group 2. Of these circRNAs, 121 were upregulated and 373 were downregulated. A total of 425 lncRNAs were differentially expressed in DR patients, of which 128 were upregulated and 297 were downregulated. One hundred and sixteen mRNAs were differentially expressed in DR patients, of which 30 were upregulated and 86 were downregulated.

**Figure 1.**
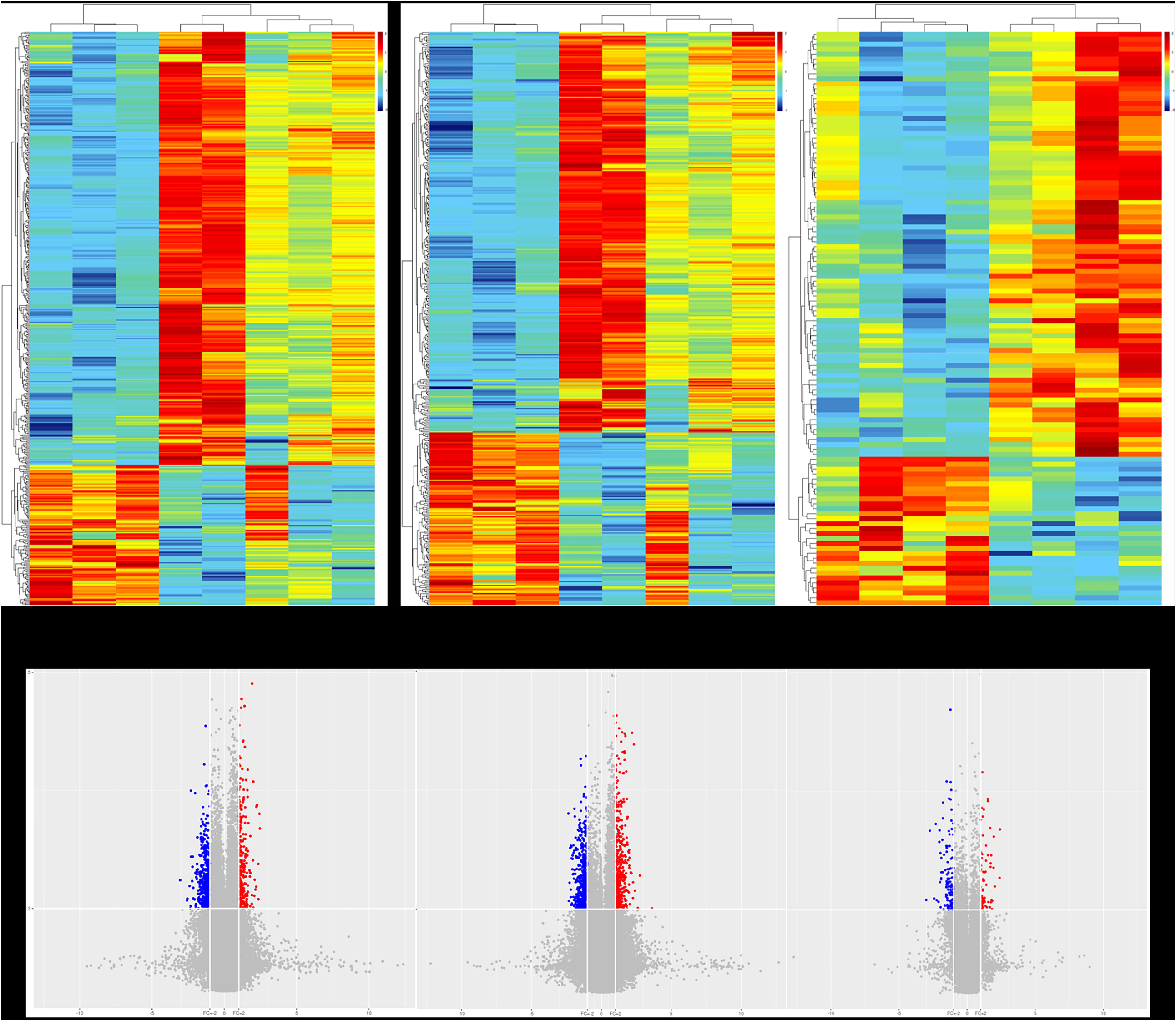
Differentially expressed circular RNAs (circRNAs), long non-coding RNAs (lncRNA)s and mRNAs in DM patients resistant to (g1) and susceptible to (g2) DR. A-C: Heatmaps show the expression of circRNAs (A), lncRNAs (B) and mRNAs (C) in different samples. D-F: Volcano plots illustrate the significantly differentially expressed circRNAs (D), lncRNAs (E) and mRNAs (F). The red dots represent the upregulated RNAs in group 1 (g1), and the blue dots represents the downregulated RNAs in group 1.

### Validation of differentially expressed circRNAs

Ten upregulated circRNAs in the microarray were used for validation. qPCR showed that 9 out of the 10 circRNAs were upregulated, which is consistent with the microarray results (Table 3).

**Table 3.**
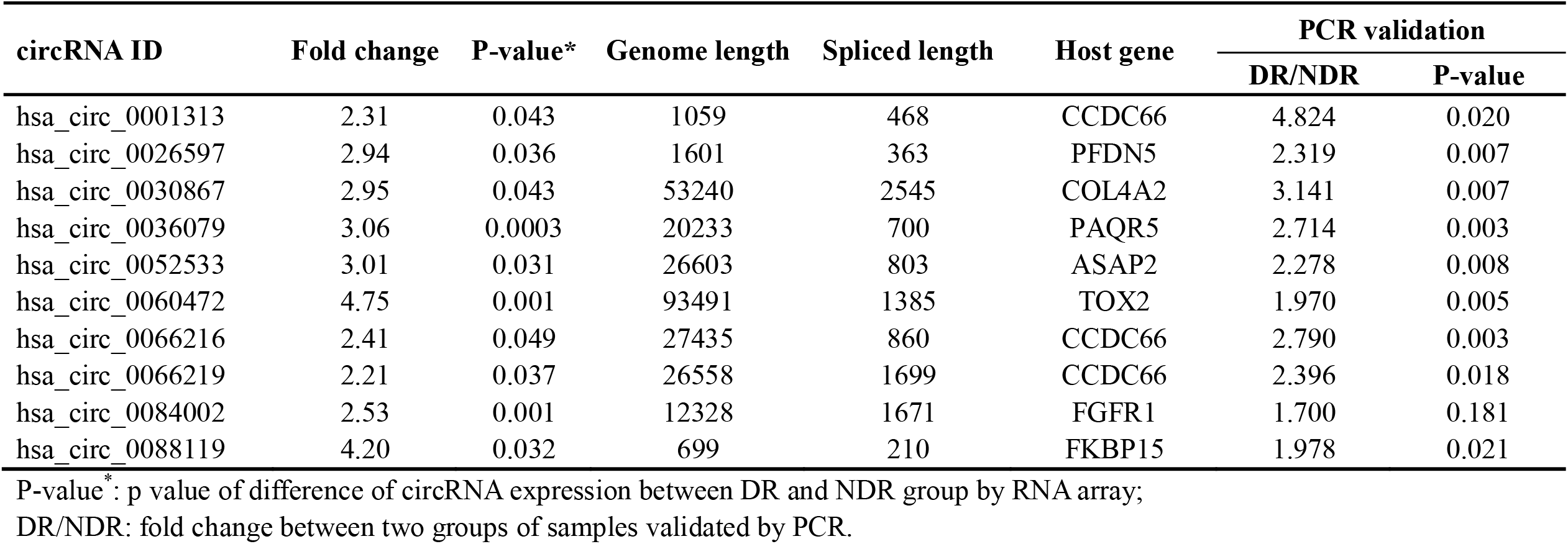
Profile of partially upregulated circRNAs identified by RNA array and results of PCR validation.

### Description of GO and pathway analysis

The results of GO enrichment analysis indicated that the differentially expressed circRNAs were involved in 24 biological processes, 14 cellular components, and 11 molecular functions. The GO analysis results of the differentially expressed circRNAs are shown in Figure 2A. The most important cellular component, molecular function, biological process were fibrillar collagen trimer, platelet-derived growth factor (PDGF) binding, and the metabolism of guanosine monophosphate (GMP), respectively. The results of the KEGG pathway analysis indicated that that the differentially expressed circRNAs were mainly associated with vitamin B6 metabolism and type 2 diabetes (Figure 2B).

**Figure 2.**
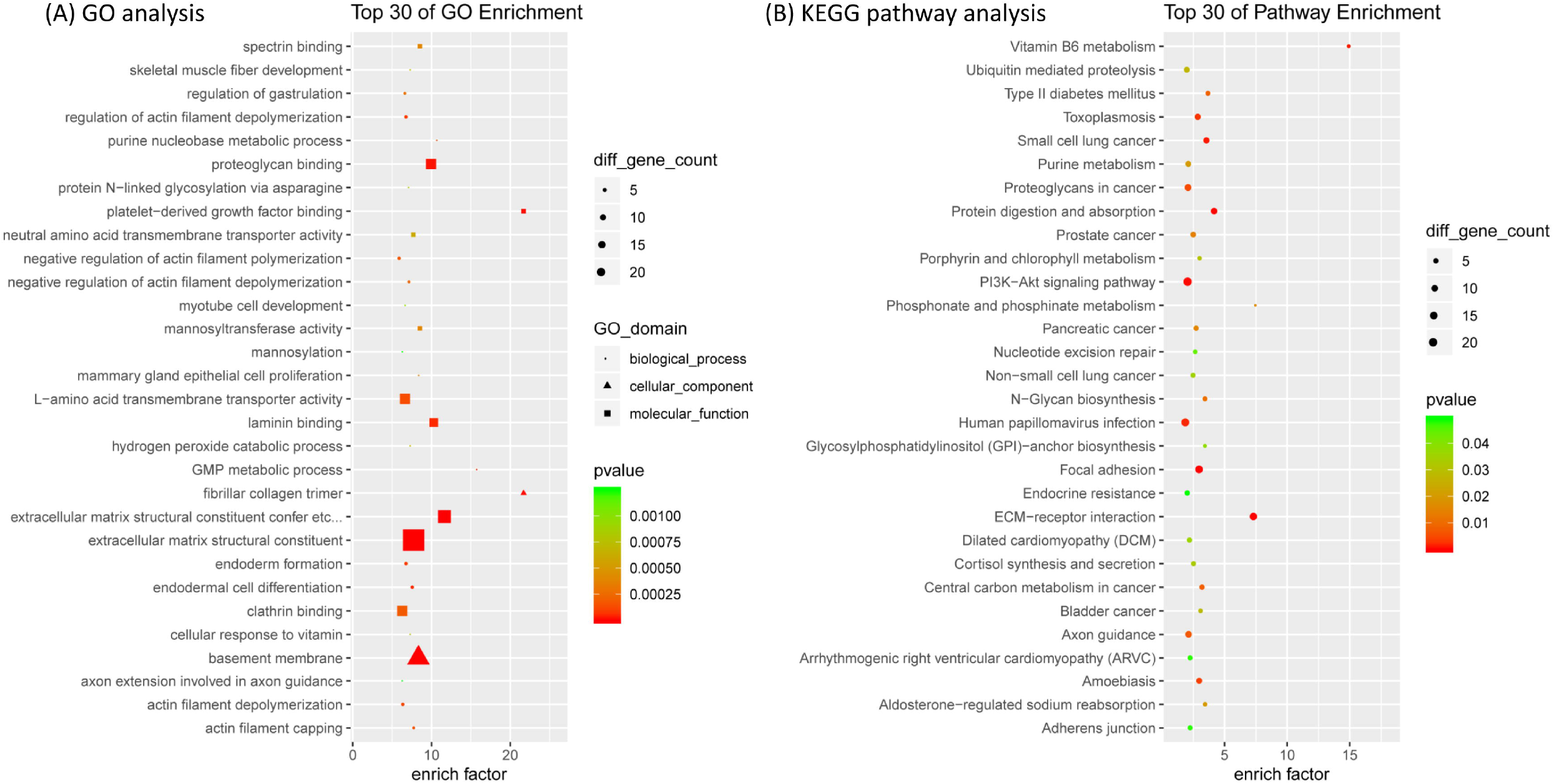
GO (A) and KEGG (B) analysis of the differentially expressed circRNAs.

The cis-target genes of the differentially expressed lncRNAs were involved in 23 biological processes, 14 cellular components, and 8 molecular functions. The GO analysis results of the cis-target genes of the differentially expressed lncRNAs are presented in Figure 3A. The biological processes include negative regulation of the Wnt signaling pathway, negative regulation of axonogenesis, and negative regulation of signaling receptor activity. The cis-target genes of lncRNAs are enriched in valine, leucine and isoleucine biosynthesis, hypoxia-inducible factor-1 (HIF-1) signaling pathway, and other pathways (Figure 3B).

**Figure 3.**
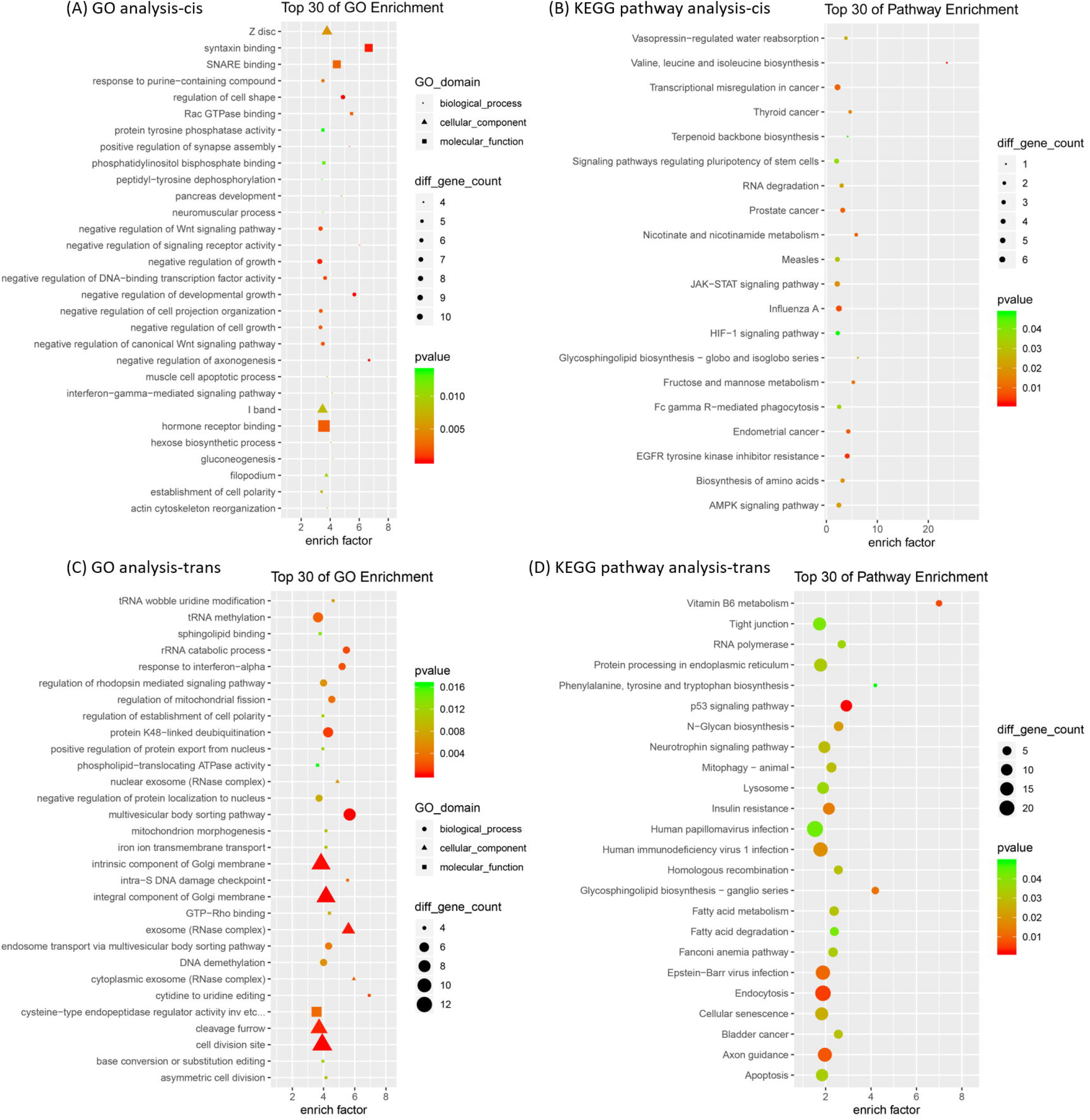
GO and KEGG analysis of the cis- and trans-target genes of the differentially expressed lncRNAs. A-B:results for cis-target genes. C-D: results for trans-target genes.

The trans-target genes were involved in 23 biological processes, 13 cellular components, and 11 molecular functions. The GO analysis results of the trans-target genes of the differentially expressed lncRNAs are shown in Figure 3C, in which biological processes include rRNA catabolic process and DNA demethylation. The trans-target genes of lncRNAs are enriched in pathways such as vitamin B6 metabolism (Figure 3D).

The differentially expressed mRNAs were involved in 20 biological processes, 13 cellular components, and 5 molecular functions. The GO analysis results of the differentially expressed mRNAs are shown in Figure 4A. The involved biological processes mainly include negative regulation of the Wnt signaling pathway and negative regulation of the apoptotic signaling pathway. KEGG analysis demonstrated that the differentially expressed mRNAs were associated with type 2 diabetes and the VEGF signaling pathway (Figure 4B).

**Figure 4.**
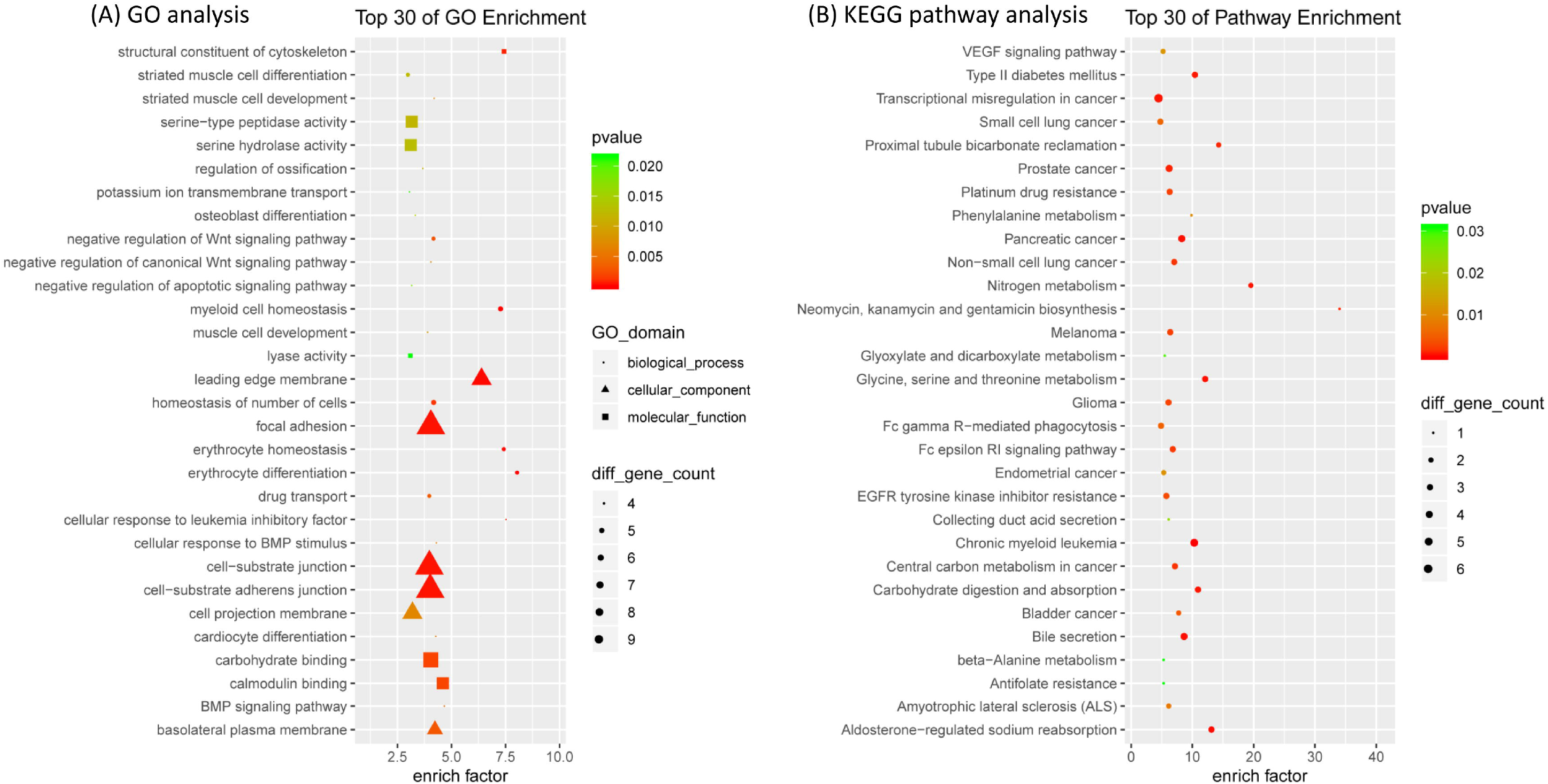
GO (A) and KEGG (B) analysis of the differentially expressed mRNAs.

### Target genes of lncRNAs and microRNAs binding to circRNAs

The cis-target genes of the top 10 up- and downregulated lncRNAs are listed in Table 4. Bioinformatic analysis was performed to predict the trans-target genes of 90 differentially expressed lncRNAs, and the prediction results contained a total of 1,445 target genes. Each differentially expressed lncRNA has 1–50 target genes. Table 5 shows the top-5 predicted miRNAs sponged by 10 circRNAs for qPCR verification. For example, hsa_circ_0001313 has five microRNA binding sites, namely hsa-miR-670-3p, hsa-miR-4326, hsa-miR-6780a-3p, hsa-miR-6885-3p, and hsa-miR-8068.

**Table 4.**
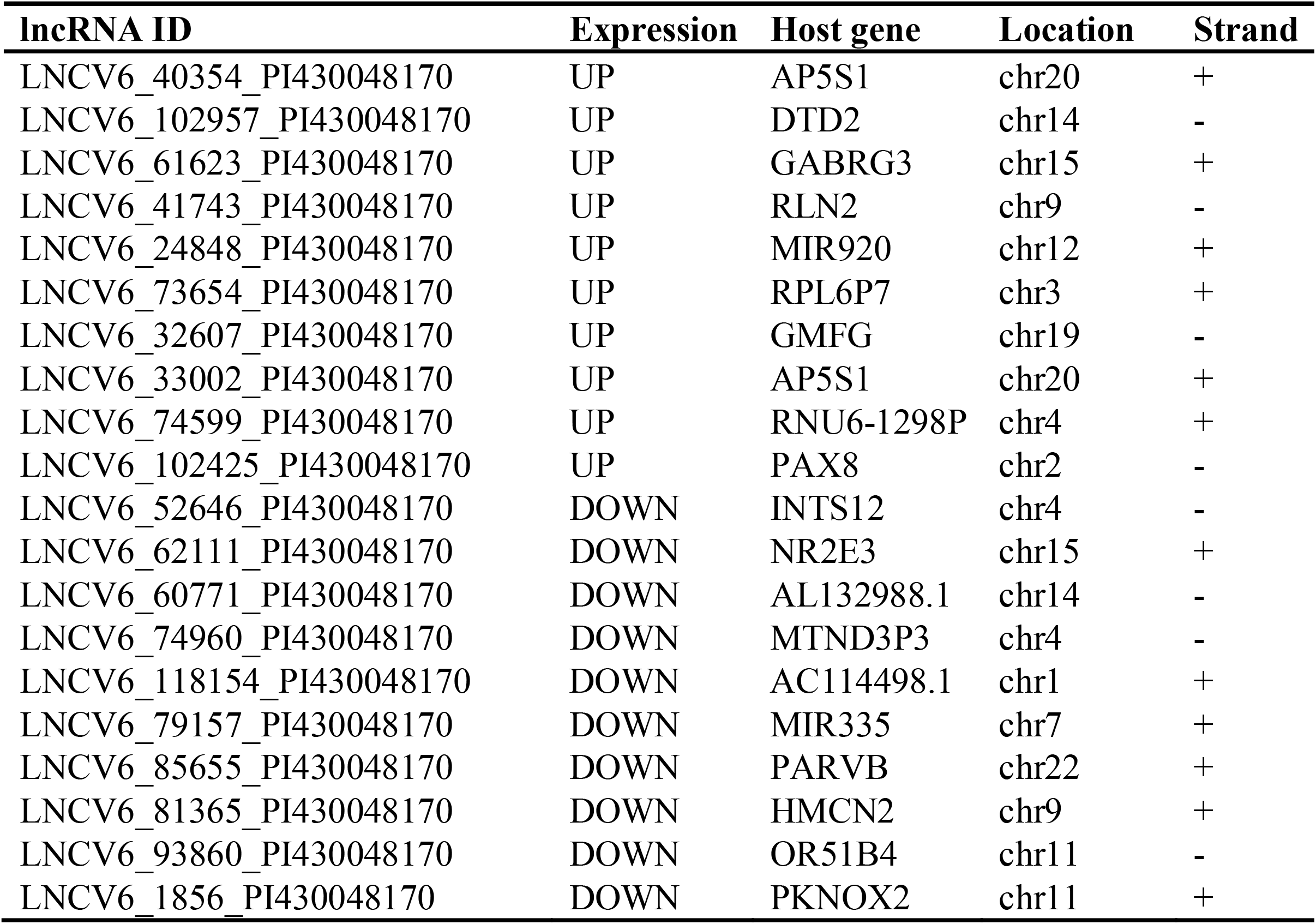
Cis target genes of top 10 upregulated and down-regulated lncRNAs

**Table 5.** Top-five predicted microRNAs sponged by the significantly unregulated circRNAs.

| **circRNA ID** | **microRNA1** | **microRNA2** | **microRNA3** | **microRNA4** | **microRNA5** |
| --- | --- | --- | --- | --- | --- |
| hsa_circ_0001313 | hsa-miR-670-3p | hsa-miR-106a-5p | hsa-miR-6780a-3p | hsa-miR-6885-3p | hsa-miR-8068 |
| hsa_circ_0066216 | hsa-miR-4676-3p | hsa-miR-452-5p | hsa-miR-892c-3p | hsa-miR-670-3p | hsa-miR-6885-3p |
| hsa_circ_0030867 | hsa-miR-6764-5p | hsa-miR-7113-5p | hsa-miR-1915-3p | hsa-miR-6783-3p | hsa-miR-1343-3p |
| hsa_circ_0066219 | hsa-miR-6875-3p | hsa-miR-578 | hsa-miR-8068 | hsa-miR-7110-3p | hsa-miR-1200 |
| hsa_circ_0088119 | hsa-miR-184 | hsa-miR-550a-3-5p | hsa-miR-550a-5p | hsa-miR-1271-3p | hsa-miR-6781-5p |
| hsa_circ_0026597 | hsa-miR-1913 | hsa-miR-324-3p | hsa-miR-4487 | hsa-miR-558 | hsa-miR-4642 |
| hsa_circ_0052533 | hsa-miR-7974 | hsa-miR-6882-3p | hsa-miR-6765-5p | hsa-miR-3615 | hsa-miR-4660 |
| hsa_circ_0060472 | hsa-miR-5787 | hsa-miR-4505 | hsa-miR-6842-3p | hsa-miR-3652 | hsa-miR-4430 |
| hsa_circ_0084002 | hsa-miR-657 | hsa-miR-4443 | hsa-miR-6515-5p | hsa-miR-4530 | hsa-miR-4274 |
| hsa_circ_0036079 | hsa-miR-665 | hsa-miR-7160-5p | hsa-miR-3605-5p | hsa-miR-5589-3p | hsa-miR-215-3p |

## Discussion

Using extreme phenotype design, we investigated the role of circRNA and lncRNA in DR development. RNA microarray showed that the expressions of circRNAs, lncRNAs, and mRNAs in the blood of patients who are susceptible to DR differ significantly from those who are immune to the disease. qPCR further verified the results. Through bioinformatics analysis, we explored the possible functions of the differentially expressed RNAs and predicted the target genes of lncRNAs and the microRNAs binding to circRNAs. The results of this study suggest that circRNAs and lncRNAs may play important regulatory roles in DR. As far as we know, this is the first study focusing on both circRNAs and lncRNAs in the development of DR by using extreme phenotype design.

In the present study, the two groups of patients with extreme phenotypes displayed significant differences in expression profiles of lncRNAs, circRNAs and mRNAs, while the traditional risk factors, such as hemoglobin A1c, cholesterol and blood pressure, were balanced between the two, which signifies potential role of the RNAs in the pathogenesis of DR. This is consistent with the conclusions of previous studies. For instance, Gu et al. analyzed expression profiles of circRNAs in the blood of healthy subjects and diabetic patients with or without retinopathy with whom no restriction on distribution of phenotype was applied, 30 circRNAs were upregulated in DR patients [13]. We previously compared the expression profiles of circRNAs in the vitreous of DR and non-diabetic patients and found that 122 circRNAs were upregulated and 9 were downregulated in the vitreous of DR patients [12]. Zhang et al. reported that 529 circRNAs were abnormally expressed in diabetic retina compared with non-diabetic retina [14]. Similarly, lncRNAs were reported to be aberrantly expressed in clinical samples of DR patients [24, 25]. The results of these studies highlighted the role of circRNAs and lncRNAs in the pathogenesis of DR. The strength of the present study is that instead of random sampling, we selected participants with extreme phenotypes from a large cohort population, functional ncRNAs are therefore enriched as much as possible, and the sensitivity to detect possible causal ncRNAs are significantly improved while the sample size is limited [26].

GO analysis supported the role of circRNAs and lncRNAs and indicated possible mechanisms in which the RNA regulators were involved. The results of GO analysis suggested that the differentially expressed circRNAs are related to PDGF binding. PDGF is an important angiogenesis regulator. It can directly induce neovascularization and fibrovascular tissue proliferation, and it plays an important role in proliferative DR [27-29]. Moreover, PDGF can regulate the distribution of tight junction proteins and change the permeability of epithelial and endothelial barriers, in addition to being related to the microangiopathy of DR [30]. Differentially expressed mRNAs and the cis-target genes of lncRNA were found to be involved in negative regulation of the Wnt signaling pathway. The Wnt signaling pathway is closely related to angiogenesis, and its abnormality is one of the important mechanisms of various ocular vascular diseases, including DR [31].

The results of the KEGG pathway analysis further supported the findings of the GO analysis, and differentially expressed lncRNAs, circRNAs, and mRNAs were found to be involved in the regulation of DR-related signaling pathways. KEGG pathway analysis suggested that differentially expressed circRNAs are associated with type 2 diabetes. In addition, vitamin B6 metabolism is the most enriched pathway for circRNAs and the trans-target genes of lncRNAs. According to the results of a national multicenter cohort study in Japan, high vitamin B6 intake can reduce the risk of DR development in patients with type 2 diabetes [32], possibly because vitamin B6 reduces lipid peroxidation and inhibits the formation of glycated hemoglobin [33]. The cis-target genes of lncRNA are enriched in the valine, leucine, and isoleucine biosynthesis pathway and the HIF-1 signaling pathway. Recent studies have demonstrated that the metabolites (metabolite markers) in the vitreous and aqueous humor of the eyes of DR patients and non-DR patients are significantly different. Pathway analysis indicated abnormalities in the valine, leucine, and isoleucine biosynthesis pathway in the eyes of DR patients [34]. Activation of the HIF-1 pathway can induce the expression of a series of vascular growth factors, such as VEGF, angiopoietin 2, and vascular endothelial-protein tyrosine phosphatase (VE-PTP), and it plays an important role in the occurrence of many ischemic ocular diseases, including DR [35]. Differentially expressed mRNAs are associated with type 2 diabetes and the VEGF signaling pathway.

In summary, lncRNAs, circRNAs, and mRNAs were differentially expressed between patients susceptible to and immune to DR. Bioinformatics analysis suggests that these RNAs may be involved in the regulation of DR-related pathways. These results suggest that lncRNAs and circRNAs are related to the pathogenesis of DR and may serve as potential biomarkers of DR.

## Data Availability

Available when appropriate request.

## Acknowledgements

We thank all participants and related staffs of this study.

## Data Availability

All relevant data are within the paper.

## References

[1] L. Liu, X. Wu, L. Liu, J. Geng, Z. Yuan, Z. Shan, L. Chen, Prevalence of diabetic retinopathy in mainland China: a meta-analysis, Plos One. 7 (2012) e45264, http://doi.org/10.1371/journal.pone.0045264.

[2] J. L. Leasher, R. R. Bourne, S. R. Flaxman, J. B. Jonas, J. Keeffe, K. Naidoo, K. Pesudovs, H. Price, R. A. White, T. Y. Wong, S. Resnikoff, H. R. Taylor, Global Estimates on the Number of People Blind or Visually Impaired by Diabetic Retinopathy: A Meta-analysis From 1990 to 2010, Diabetes Care. 39 (2016) 1643–1649, http://doi.org/10.2337/dc15-2171.

[3] J. W. Yau, S. L. Rogers, R. Kawasaki, E. L. Lamoureux, J. W. Kowalski, T. Bek, S. J. Chen, J. M. Dekker, A. Fletcher, J. Grauslund, S. Haffner, R. F. Hamman, M. K. Ikram, T. Kayama, B. E. Klein, R. Klein, S. Krishnaiah, K. Mayurasakorn, J. P. O’Hare, T. J. Orchard, M. Porta, M. Rema, M. S. Roy, T. Sharma, J. Shaw, H. Taylor, J. M. Tielsch, R. Varma, J. J. Wang, N. Wang, S. West, L. Xu, M. Yasuda, X. Zhang, P. Mitchell, T. Y. Wong, Global prevalence and major risk factors of diabetic retinopathy, Diabetes Care. 35 (2012) 556–564, http://doi.org/10.2337/dc11-1909.

[4] Diabetes in China: mapping the road ahead, Lancet Diabetes Endocrinol. 2 (2014) 923, http://doi.org/10.1016/S2213-8587(14)70189-5.

[5] L. Wang, P. Gao, M. Zhang, Z. Huang, D. Zhang, Q. Deng, Y. Li, Z. Zhao, X. Qin, D. Jin, M. Zhou, X. Tang, Y. Hu, L. Wang, Prevalence and Ethnic Pattern of Diabetes and Prediabetes in China in 2013, JAMA. 317 (2017) 2515–2523, http://doi.org/10.1001/jama.2017.7596.

[6] J. M. Lachin, S. Genuth, D. M. Nathan, B. Zinman, B. N. Rutledge, Effect of glycemic exposure on the risk of microvascular complications in the diabetes control and complications trial--revisited, Diabetes. 57 (2008) 995–1001, http://doi.org/10.2337/db07-1618.

[7] R. Klein, B. E. Klein, S. E. Moss, K. J. Cruickshanks, The Wisconsin Epidemiologic Study of Diabetic Retinopathy: XVII. The 14-year incidence and progression of diabetic retinopathy and associated risk factors in type 1 diabetes, Ophthalmology. 105 (1998) 1801–1815, http://doi.org/10.1016/S0161-6420(98)91020-X.

[8] N. H. Arar, B. I. Freedman, S. G. Adler, S. K. Iyengar, E. Y. Chew, M. D. Davis, S. G. Satko, D. W. Bowden, R. Duggirala, R. C. Elston, X. Guo, R. L. Hanson, R. P. J. Igo, E. Ipp, P. L. Kimmel, W. C. Knowler, J. Molineros, R. G. Nelson, M. V. Pahl, S. R. E. Quade, R. S. Rasooly, J. I. Rotter, M. F. Saad, M. Scavini, J. R. Schelling, J. R. Sedor, V. O. Shah, P. G. Zager, H. E. Abboud, Heritability of the Severity of Diabetic Retinopathy: The FIND-Eye Study, Invest Ophth Vis Sci. 49 (2008) 3839–3845, http://doi.org/10.1167/iovs.07-1633.

[9] F. Kopp, J. T. Mendell, Functional Classification and Experimental Dissection of Long Noncoding RNAs, Cell. 172 (2018) 393–407, http://doi.org/10.1016/j.cell.2018.01.011.

[10] J. D. Ransohoff, Y. Wei, P. A. Khavari, The functions and unique features of long intergenic non-coding RNA, Nat Rev Mol Cell Biol. 19 (2018) 143–157, http://doi.org/10.1038/nrm.2017.104.

[11] B. Han, J. Chao, H. Yao, Circular RNA and its mechanisms in disease: From the bench to the clinic, Pharmacol Ther. 187 (2018) 31–44, http://doi.org/10.1016/j.pharmthera.2018.01.010.

[12] M. He, W. Wang, H. Yu, D. Wang, D. Cao, Y. Zeng, Q. Wu, P. Zhong, Z. Cheng, Y. Hu, L. Zhang, Comparison of expression profiling of circular RNAs in vitreous humour between diabetic retinopathy and non-diabetes mellitus patients, Acta Diabetol. (2019), http://doi.org/10.1007/s00592-019-01448-w.

[13] Y. Gu, G. Ke, L. Wang, E. Zhou, K. Zhu, Y. Wei, Altered Expression Profile of Circular RNAs in the Serum of Patients with Diabetic Retinopathy Revealed by Microarray, Ophthalmic Res. 58 (2017) 176–184, http://doi.org/10.1159/000479156.

[14] S. J. Zhang, X. Chen, C. P. Li, X. M. Li, C. Liu, B. H. Liu, K. Shan, Q. Jiang, C. Zhao, B. Yan, Identification and Characterization of Circular RNAs as a New Class of Putative Biomarkers in Diabetes Retinopathy, Invest Ophthalmol Vis Sci. 58 (2017) 6500–6509, http://doi.org/10.1167/iovs.17-22698.

[15] O. Wapinski, H. Y. Chang, Long noncoding RNAs and human disease, Trends Cell Biol. 21 (2011) 354–361, http://doi.org/10.1016/j.tcb.2011.04.001.

[16] Q. Gong, G. Su, Roles of miRNAs and long noncoding RNAs in the progression of diabetic retinopathy, Biosci Rep. 37 (2017), http://doi.org/10.1042/BSR20171157.

[17] B. Yan, Z. F. Tao, X. M. Li, H. Zhang, J. Yao, Q. Jiang, Aberrant expression of long noncoding RNAs in early diabetic retinopathy, Invest Ophthalmol Vis Sci. 55 (2014) 941–951, http://doi.org/10.1167/iovs.13-13221.

[18] Grading diabetic retinopathy from stereoscopic color fundus photographs--an extension of the modified Airlie House classification. ETDRS report number 10. Early Treatment Diabetic Retinopathy Study Research Group, Ophthalmology. 98 (1991) 786–806,

[19] The Gene Ontology (GO) project in 2006, Nucleic Acids Res. 34 (2006) D322–D326, http://doi.org/10.1093/nar/gkj021.

[20] A. Y. Gracey, E. J. Fraser, W. Li, Y. Fang, R. R. Taylor, J. Rogers, A. Brass, A. R. Cossins, Coping with cold: An integrative, multitissue analysis of the transcriptome of a poikilothermic vertebrate, Proc Natl Acad Sci U S A. 101 (2004) 16970–16975, http://doi.org/10.1073/pnas.0403627101.

[21] M. Ashburner, C. A. Ball, J. A. Blake, D. Botstein, H. Butler, J. M. Cherry, A. P. Davis, K. Dolinski, S. S. Dwight, J. T. Eppig, M. A. Harris, D. P. Hill, L. Issel-Tarver, A. Kasarskis, S. Lewis, J. C. Matese, J. E. Richardson, M. Ringwald, G. M. Rubin, G. Sherlock, Gene ontology: tool for the unification of biology. The Gene Ontology Consortium, Nat Genet. 25 (2000) 25–29, http://doi.org/10.1038/75556.

[22] S. Draghici, P. Khatri, A. L. Tarca, K. Amin, A. Done, C. Voichita, C. Georgescu, R. Romero, A systems biology approach for pathway level analysis, Genome Res. 17 (2007) 1537–1545, http://doi.org/10.1101/gr.6202607.

[23] M. Kanehisa, The KEGG resource for deciphering the genome, Nucleic Acids Res. 32 (2004) 277D–280D, http://doi.org/10.1093/nar/gkh063.

[24] L. Yin, Z. Sun, Q. Ren, X. Su, D. Zhang, Long Non-Coding RNA BANCR Is Overexpressed in Patients with Diabetic Retinopathy and Promotes Apoptosis of Retinal Pigment Epithelial Cells, Med Sci Monit. 25 (2019) 2845–2851, http://doi.org/10.12659/MSM.913359.

[25] Q. Li, L. Pang, W. Yang, X. Liu, G. Su, Y. Dong, Long Non-Coding RNA of Myocardial Infarction Associated Transcript (LncRNA-MIAT) Promotes Diabetic Retinopathy by Upregulating Transforming Growth Factor-beta1 (TGF-beta1) Signaling, Med Sci Monit. 24 (2018) 9497–9503, http://doi.org/10.12659/MSM.911787.

[26] I. J. Barnett, S. Lee, X. Lin, Detecting rare variant effects using extreme phenotype sampling in sequencing association studies, Genet Epidemiol. 37 (2013) 142–151, http://doi.org/10.1002/gepi.21699.

[27] K. Mori, P. Gehlbach, A. Ando, G. Dyer, E. Lipinsky, A. G. Chaudhry, S. F. Hackett, P. A. Campochiaro, Retina-specific expression of PDGF-B versus PDGF-A: vascular versus nonvascular proliferative retinopathy, Invest Ophthalmol Vis Sci. 43 (2002) 2001–2006,

[28] P. A. Campochiaro, S. F. Hackett, S. A. Vinores, J. Freund, C. Csaky, W. LaRochelle, J. Henderer, M. Johnson, I. R. Rodriguez, Z. Friedman, A. Et, Platelet-derived growth factor is an autocrine growth stimulator in retinal pigmented epithelial cells, J Cell Sci. 107 (Pt 9) (1994) 2459–2469,

[29] S. D. Schoenberger, S. J. Kim, R. Shah, J. Sheng, E. Cherney, Reduction of interleukin 8 and platelet-derived growth factor levels by topical ketorolac, 0.45%, in patients with diabetic retinopathy, Jama Ophthalmol. 132 (2014) 32–37, http://doi.org/10.1001/jamaophthalmol.2013.6203.

[30] N. S. Harhaj, A. J. Barber, D. A. Antonetti, Platelet-derived growth factor mediates tight junction redistribution and increases permeability in MDCK cells, J Cell Physiol. 193 (2002) 349–364, http://doi.org/10.1002/jcp.10183.

[31] Z. Wang, C. H. Liu, S. Huang, J. Chen, Wnt Signaling in vascular eye diseases, Prog Retin Eye Res. 70 (2019) 110–133, http://doi.org/10.1016/j.preteyeres.2018.11.008.

[32] C. Horikawa, R. Aida, C. Kamada, K. Fujihara, S. Tanaka, S. Tanaka, A. Araki, Y. Yoshimura, T. Moriya, Y. Akanuma, H. Sone, Vitamin B6 intake and incidence of diabetic retinopathy in Japanese patients with type 2 diabetes: analysis of data from the Japan Diabetes Complications Study (JDCS), Eur J Nutr. (2019), http://doi.org/10.1007/s00394-019-02014-4.

[33] S. K. Jain, G. Lim, Pyridoxine and pyridoxamine inhibits superoxide radicals and prevents lipid peroxidation, protein glycosylation, and (Na+ + K+)-ATPase activity reduction in high glucose-treated human erythrocytes, Free Radic Biol Med. 30 (2001) 232–237, http://doi.org/10.1016/s0891-5849(00)00462-7.

[34] H. Wang, J. Fang, F. Chen, Q. Sun, X. Xu, S. H. Lin, K. Liu, Metabolomic profile of diabetic retinopathy: a GC-TOFMS-based approach using vitreous and aqueous humor, Acta Diabetol. 57 (2020) 41–51, http://doi.org/10.1007/s00592-019-01363-0.

[35] P. A. Campochiaro, Molecular pathogenesis of retinal and choroidal vascular diseases, Prog Retin Eye Res. 49 (2015) 67–81, http://doi.org/10.1016/j.preteyeres.2015.06.002.

